# Evaluating Magnetic Stimulation as an Innovative Approach for Treating Dry Eye Syndrome: Safety and Efficacy Initial study

**DOI:** 10.1101/2024.03.27.24304988

**Authors:** Hadas Ben-Eli, Shimon Perelman, Denise Wajnsztajn, Abraham Solomon

## Abstract

**Objective:** The objective of this study was to assess the safety and preliminary efficacy of repetitive magnetic stimulation (RMS) as an intervention for dry eye syndrome, focusing on symptom reduction.

**Methodology:** This investigation involved 22 adult participants diagnosed with moderate to severe dry eye syndrome. These individuals were subjected to RMS treatment targeting one or both eyes using the VIVEYE - Ocular Magnetic Neurostimulation System Ver 1.0 (Epitech-Mag LTD; NIH clinical trials registry #NCT03012698). A placebo-controlled group was also included for comparative analysis, with all subjects being monitored over a three-month period. The evaluation of safety encompassed monitoring changes in best corrected visual acuity, ocular pathology, and reporting of adverse events. Participant tolerance was gauged through questionnaires, measurements of intraocular pressure (IOP), Schirmer’s test, and vital signs. The efficacy of the treatment was assessed by comparing pre- and post-treatment scores on fluorescein staining (according to the National Eye Institute (NEI) grading) and patient-reported outcomes.

**Results:** The study found no significant changes in visual acuity, IOP, or Schirmer’s test results between the RMS-treated and control groups (p<0.05), indicating RMS does not adversely affect these ocular functions. However, RMS treatment was associated with improved tear film stability (p=0.198 vs. p=0.045) and corneal health (p=0.52 vs. p=0.004), with no improvements in the control group. Initial symptom improvement was observed in both RMS-treated and placebo groups (p=0.007 vs p=0.008), suggesting RMS’s potential for treating ocular surface conditions.

**Conclusion:** The findings of this study introduce repetitive magnetic stimulation (RMS) as a promising therapeutic option for dry eye syndrome, demonstrating its capability to promote corneal epithelium repair, enhance tear film stability, and improve subjective symptom evaluations without adversely affecting intraocular pressure, visual acuity, or tear production. This confirms the safety and suggests the efficacy of RMS therapy for dry eye conditions.

## Introduction

Dry Eye Disease (DED), also referred to as keratoconjunctivitis sicca (KCS), is identified by the International Dry Eye Workshop (DEWS, 2007) as a multifactorial disorder impacting the tears and ocular surface. This chronic condition leads to discomfort, visual disturbances, an unstable tear film, and possible ocular surface damage [1]. DED is a common diagnosis in ophthalmology, with a growing prevalence ranging from 5% to 33% in the adult population worldwide [1–3], and even higher, up to 87%, in visual display terminal workers [4]. Notably, 78% of DED patients are women [2]. The symptoms, including irritation, stinging, and fluctuating visual disturbances, can progress to severe complications like vision impairment and corneal damage if left untreated [5], [6].

It is assumed that all the intrinsic and extrinsic etiology and risk factors that initiate DED development are associated with the disruption of the structure or function of one or more of the tear film layers, which leads to Corneo-conjunctival epithelial damage[1], [2]. Since the cornea is one of the most highly innervated tissues in the body, with terminal nerves ending in superficial layers of the epithelium in close contact with the environment, a wide spectrum of stimuli trigger the trigeminal pathway to the somatosensory cortex and limbic system, resulting in the sensation of pain[3].

Traditional treatments largely involve topical lubricants and anti-inflammatory drops. Despite their widespread use, these methods pose economic challenges and often have limited efficacy, particularly in severe cases, necessitating long-term use [3], [9], [10]. Recent advancements have introduced treatments based on heating and massaging the eyelids and meibomian glands, but these can be painful and provide only short-term relief [11–13]. As of today there are no therapeutic options that are significantly useful in the treatment of dry eye disorders. A novel non-invasive treatment based on repetitive magnetic stimulation (RMS) was recently studied in a pre-clinical trial and was reported to be extremely useful in protection of the corneal epithelium in short-term exposure keratopathy in a rabbit model[4]. Based on these preliminary results, a first-in-human study was performed by our group. The current study was the first clinical trial using this method of treatment on human subjects with DED. Neuromodulation is a recent approach that utilizes electrical signals to modulate abnormal neural function through neurostimulation. This newly developed technique stimulates the nervous system via electrical currents[5], either by external electric field[4] or intranasal neurostimulation therapy, to stimulate tear production[5], [6]. The RMS treatment was developed based on the transcranial magnetic stimulation (TMS) approach, which is based on neurostimulation and neuromodulation and is in clinical use (FDA approved since 2008) for treatment of a variety of neurological and psychiatric disorders such as obsessive-compulsive disorder (OCD)[7], depressive disorders[8], schizophrenia, and Parkinson’s disease[9]. Neuromodulation, as defined by the International Neuromodulation Society, involves altering nerve activity through stimuli like electrical stimulation or chemical agents to a specific neurological site [17]. Repetitive transcranial magnetic stimulation (rTMS) is thought to modulate neuronal systems through various mechanisms. These include altering neurotransmitter and ion channel activities, inducing intra-cortical inhibition and long-term potentiation, and affecting gene expression and growth factor production. It also impacts signaling pathways and the glutamate-mediated blood-brain barrier. Additionally, rTMS is believed to stimulate parasympathetic innervation to the lacrimal glands [11][12]. The current study focuses on the application of RMS in humans, following its efficacy in decreasing epithelial corneal erosions in a rabbit model of exposure keratopathy [4]. This study aims to assess the safety and efficacy of a novel non-invasive instrument designed for treating Dry Eye Disease (DED) patients, marking the first human trial of the RMS procedure.

## Methods

### Study design

Prospective, hospital-based, interventional, open-label, single group assignment study (#ClinicalTrials.gov Identifier: NCT03012698)[10].

#### Study aims

The primary objective of this study was to evaluate the safety of repetitive magnetic stimulation (RMS) as a treatment for dry eye disease. The secondary objectives were to assess the tolerability of the treatment and to determine its preliminary efficacy in reducing signs and symptoms of dry eye.

### Study endpoints

The primary endpoint of this study was the evaluation of successful RMS treatment for dry eye disease measured by a lack of deterioration in the best corrected visual acuity (BCVA) and reduction in DED signs and symptoms. The secondary endpoints were based on safety and tolerability measures: pathological ocular changes observed in a slit lamp bio microscopy assessment, any adverse events, questionnaire-based tolerability assessment, intraocular pressure, Schirmer’s test, and vital signs (heart rate, blood pressure and body temperature) are all indications for trial termination. Efficacy secondary endpoints include clinically and statistically significant reduced fluorescein staining scores between baseline and post-treatment visits at any of the follow-up time point, reduced ocular discomfort between baseline and post treatment visits at any of follow up time points (questionnaire score) and an improvement in tear film as measured in Tear Break out Time (TBUT).

### Participants

This study included 22 male and female adult patients with moderate to severe dry eye syndrome classified by severity of signs and symptoms[11] with different etiologies (meibomian gland Dysfunction (MGD), Sjogren’s Syndrome (SS), aqueous tear deficiency (ATD) and graft versus host disease (GVHD)), recruited at the Ophthalmology Clinics of Hadassah Medical Center. Patients received one-time treatment with the VIVEYE - Ocular Magnetic Neurostimulation System Ver 1.0 (Epitech-Mag LTD., Israel, 2016).

The follow up period was 12 weeks and involved 10 evaluations (screening, baseline, treatment, 1 day, 1 month, 2 months, 3 months, and bi-weekly phone calls). At each visit, patients were examined for treatment safety and efficacy, as well as symptomatic grading, as will be detailed. Contact lens wearers were asked to refrain from using contact lenses for the duration of the study.

The study was approved by the national Ministry of Health (#20162621) and by the institutional Helsinki committees of Hadassah Medical Center ( #HMO-0405-19).This study was also registered in the NIH clinical trials registry (#ClinicalTrials.gov Identifier: NCT03012698 [10]). Recruitment started on December 4, 2017 and ended on April 25, 2021. All participants signed a consent form prior to enrollment after receiving a verbal and written explanation on the study, and all data was coded and analyzed anonymously.

#### Inclusion / exclusion criteria

This study included males and females between the ages of 18-80 years, who have been diagnosed with moderate to severe dry eyes and were able to understand the requirements of the study protocol and providing informed consent. Individuals with other ocular surface pathologies requiring treatment beyond ocular lubricants and conventional eyelid hygiene, concurrent ocular diseases such as ocular infection or pterygium, recent ocular surgery (within the last 6 months) or LASIK (within the past year prior the recruitment), ocular injury or ocular herpes infection within the last 3 months were excluded. Also, patients who have recently taken central nervous system drugs, require contact lenses during the study, have thyroid disorders, alcoholism, are pregnant or nursing, have HIV, or have various implants like pacemakers or cochlear implants. Additionally, patients with significant heart or brain diseases, a history of neurological conditions, or those who have recently participated in another ophthalmic trial are also excluded.

#### Instrument

The VIVEYE - Ocular Magnetic Neurostimulation System Ver 1.0 (Epitech-Mag LTD., Israel, 2016) is a non-invasive stimulation device intended for the application of localized electromagnetic stimulation to the cornea in adult patients with dry eye disorders (device overview at Supplementary Table S1). Its main components are a Rapid2 stimulator unit and a pair of coil applicators. The applicators are attached to a positioning device for adjustment of their position relative to the patient’s eyes. At any given time, only one coil can be connected to the stimulator so only one eye is treated. The system uses a commercial ophthalmic table (CE marked) and chin rest to adjust to various patient sizes. The Rapid2 stimulator is the central component of the system and controls the various properties of the magnetic stimulation, such as intensity and rate. It consists of a generator unit, a touchscreen for selecting the treatment parameters and triggering the stimulation, and a set of coils. This device was approved and standardized by the international harmonized standards for clinical investigation of medical devices (ISO 14155, Clinical investigation of medical devices for human subjects).

Each eye is treated separately in the VIVEYE - Ocular Magnetic Neurostimulation System, taking about 11 minutes to complete a set of magnetic pulses. Each participant received demo magnetic pulses on the hand and the eye prior the treatment to understand the sensation of the light vibration stimulated by the device. Any metallic objects around the face were removed before treatment. Each participant completed a set of 32 magnetic pulses in gradually increasing power, reaching a maximal intensity of 45%.

#### Safety and efficacy tests

Safety and efficacy of the VIVEYE - Ocular Magnetic Neurostimulation System was based on ophthalmic and vital signs that were evaluated at each follow up visit. Safety tests included assessment of treatment-related adverse or serious adverse events, best corrected visual acuity (BCVA, ETDRS chart, LogMAR units), intra-ocular pressure (IOP) measurement, Schirmer II test (with local anesthesia, measuring mm/5 min)[12], external eye examination by slit lamp biomicroscopy assessment, fundus examination (with dilation at baseline visit, 1 day, 1 week and 12 weeks post treatment) and SD-OCT[13] at baseline visit, 1 week and 12 weeks post treatment.

Treatment efficacy tests included TBUT[14], [15] which was repeated three times with the mean result recorded, and corneal fluorescein staining photography (BI 900)), in which two strips of fluorescein were diluted in 500µl of saline solution for 1.5 minutes, and then inserted of 0.2µl to the conjunctival fornix. Subjective grading of the corneal erosion and fluorescein staining was done using the well-validated National Eye Institute (NEI) grading scale, commonly used in clinical settings[16], [17]. The NEI scale divides the cornea into five different areas, and each era is given a subjective score between 0 and 3 based on the number, size and confluence of the superficial punctate erosions. To fully evaluate the efficacy for the novel approach for treating Dry Eye Disease, we employed a comprehensive approach to understand the full impact of our intervention on participants’ symptoms and quality of life. For this purpose, we selected two distinct questionnaires, each with a unique focus and strength, to ensure a comprehensive evaluation. At each visit the patients answered Quality of life (QoL) and eye dryness symptoms questionnaires (modified SPEED questionnaire [18] and PROWL questionnaire [19]) and the visual analog scale (VAS) for eye dryness [20] and monitoring of use of ophthalmic lubricants. Quality of Life (QOL) surveys were assessed for each participant throughout the three-month follow-up duration. To normalize the outcomes, a scoring system was applied where the minimum QOL scores were allocated a value of 1, and the maximum scores were given a value of 5. This method of scoring was crucial for integrating the results from the different QoL questionnaires used in our study, allowing for a unified analysis of the data.

#### Course of experiment

Patients consulting the cornea clinics at Hadassah Medical Center with complaints of dry eyes were approached for participation. Those who potentially qualified to participate in the study underwent a screening test to ensure they met the inclusion criteria and to grade the severity of eye dryness with a validated scaling approach of signs and symptoms[11] Only patients with moderate to severe levels of eye dryness were included in the current study. The severity grading and all the ophthalmic clinical evaluations were done by cornea specialists (A.S, D.W). A detailed systemic and ophthalmic history was documented for each participant.

The follow-up period was 12 weeks and included 8 visits: screening, baseline, treatment, 1 day, 1 week, 4 weeks, 8 weeks and 12 weeks post-treatment, as well as bi-weekly phone calls at 2, 6 and 10 weeks (see study visit scheme at Supplementary Table S2). Each follow-up by phone included an eye dryness questionnaire and drug intake monitoring. Follow up visits in the clinic also included many safety and efficacy tests, as was detailed above. The study was divided into two phases. In the first phase, therapy was administered to only one eye per patient, leaving the other eye with no treatment (n=7), and patients were not informed about which eye was treated. Although both eyes underwent identical procedures, only one eye received magnetic stimulation, while the other eye received a placebo treatment. In the following phase, the participants were divided into two groups: one received treatment for both eyes during the same session (n=9), whereas the other group received placebo treatments for both eyes (n=6). Several participants failed to complete all the planned follow-up visits, resulting in their exclusion from the study’s statistical evaluation (n=2).

#### Statistical analysis

Quantitative variables are presented as mean and standard deviation, and changes across different time points were assessed using the Friedman nonparametric test. Comparison between two independent groups’ quantitative variables was conducted using either the t-test or the non-parametric Mann-Whitney (M-W) test. Simultaneous evaluation of time, treatment effects, and their interaction was achieved through the application of the repeated measures ANOVA model, employing the greenhouse Geiser test. Associations between two categorical variables were tested using the Chi-square test and Fisher’s exact test. The utilization of nonparametric tests was prompted by the limited sample size. All statistical tests were two-tailed, with a significance threshold set at p-value ≤ 0.05 to determine statistical significance. Statistical analysis was performed using JMP® Statistical Discovery software, version 14.3.0 from SAS® Institute Inc., Cary NC.

## Results

A total of 20 patients were included in this study, with mean age [and SD] of 51.4 [18.6] for the treated group and 47.7 [17.9] for the non-treated group. The overall age range was 22-79 (Table 1). As expected, most of the study participants were women in both groups (N =16; 84% and N =22; 88% respectively.) Sjogern was the most common clinical manifestation among the treated and non-treated group. Other common etiologies were aqueous tear deficiency and meibomian gland disfunction. While composing mostly of these 3 etiologies, there was no statistically significant differences in the dispersion of different etiologies between the treatment groups. [Table 1]. Baseline measurements of Intra-Ocular Pressure (IOP), Best Corrected Visual Acuity (BCVA) and Shirmer test exhibited no comparable values between the two groups.

**Table 1.**
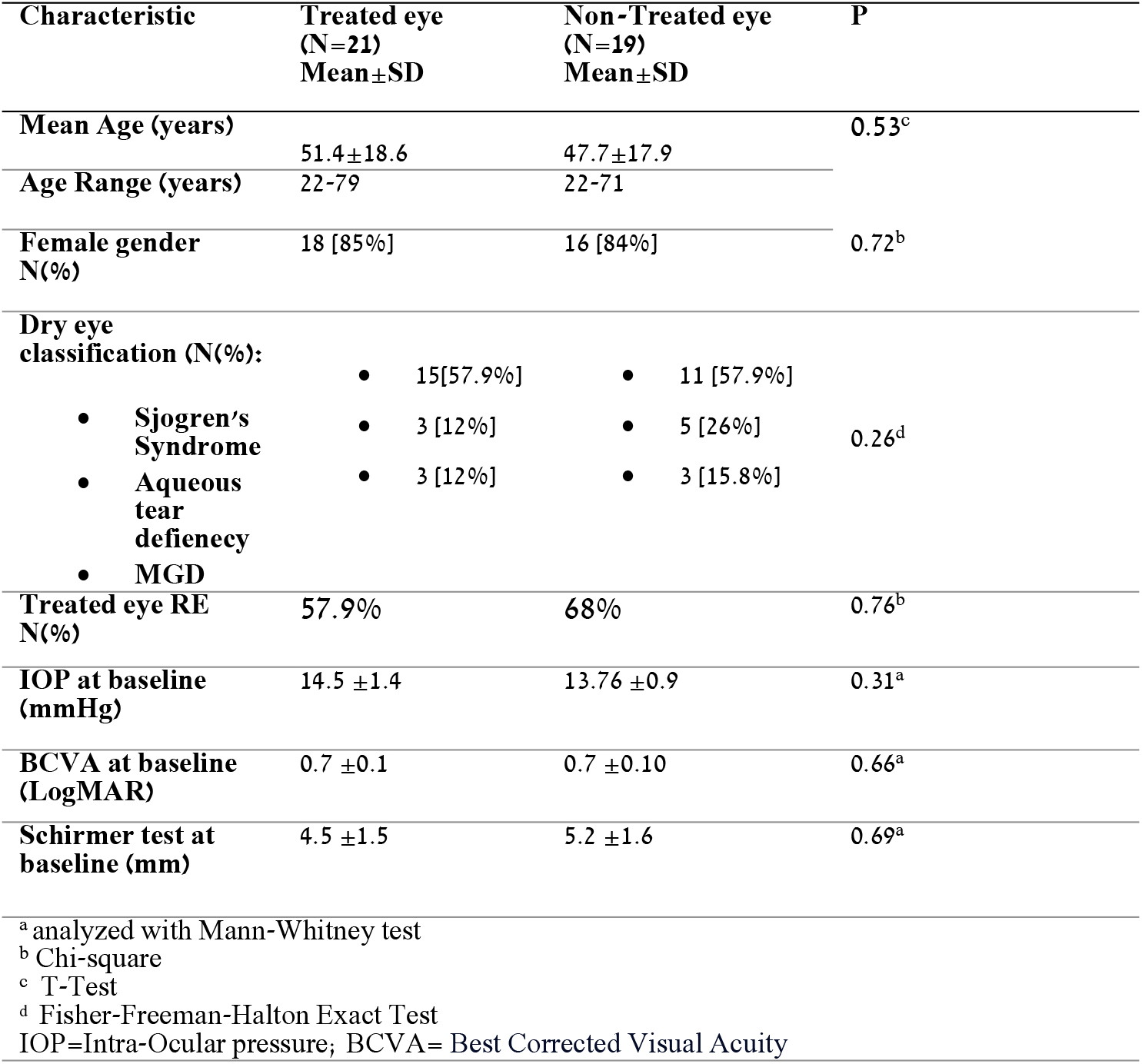
Basic characteristics study participants.

We found no significant differences in baseline intraocular pressure, best-corrected visual acuity, or tear production between the group that received treatment and the control group [Table 1].

The safety of RMS was examined using various tests. One of them was Best corrected visual acuity (BCVA) over several time points of both treated and non-treated eyes were compared and presented at [Table 2]. The BCVA measurements were obtained for both treated and non-treated eyes at enrolment and subsequently at 1 week, 1 month, 2-month and 3-month intervals. The mean BCVA values demonstrated no statistically significant differences between the treated and non-treated eyes across different time points as shown in [Table 2].

**Table 2:** Safety of Repetitive Magnetic Stimulation.

| Treatment | Measurement | Time | Mean ± SD | P Value of Change Over Time |
| --- | --- | --- | --- | --- |
| **Treated**<br>**N=21** | **BCVA** | Enrolment | 0.22±0.28 | 0.13<sup>a</sup> |
| | | 1 Week | 0.17±0.26 | |
| | | 1 Month | 0.15±0.26 | |
| | | 2 Months | 0.15±0.26 | |
| | | 3 Months | 0.19±0.26 | |
| | **IOP** | Enrolment | 13.76±2.57 | 0.16<sup>a</sup> |
| | | 1 Week | 12.40±2.68 | |
| | | 1 Month | 11.96±3.43 | |
| | | 2 Months | 11.96±2.65 | |
| | | 3 Months | 12.90±3.13 | |
| | **Schirmer** | Enrolment | 5.24±4.32 | 0.42<sup>b</sup> |
| | | 1 Week | 4.44±3.65 | |
| | | 1 Month | 4.64±4.00 | |
| | | 2 Months | 4.13±3.70 | |
| | | 3 Months | 3.62±2.71 | |
| **Not Treated**<br>**N=19** | **BCVA** | Enrolment | 0.14±0.27 | 0.39<sup>a</sup> |
| | | 1 Week | 0.15±0.28 | |
| | | 1 Month | 0.14±0.26 | |
| | | 2 Months | 0.14±0.26 | |
| | | 3 Months | 0.16±0.28 | |
| | **IOP** | Enrolment | 14.53±3.24 | 0.74<sup>a</sup> |
| | | 1 Week | 13.05±3.31 | |
| | | 1 Month | 13.84±3.43 | |
| | | 2 Months | 13.42±3.31 | |
| | | 3 Months | 13.89±3.13 | |
| | **Schirmer** | Enrolment | 4.53±3.65 | 0.51<sup>b</sup> |
| | | 1 Week | 3.89±2.76 | |
| | | 1 Month | 3.47±2.95 | |
| | | 2 Months | 4.5±4.1 | |
| | | 3 Months | 4.47±3.60 | |

<sup>a</sup> Calculated using Freidman Test<sup>b</sup> Calculated using Greenhouse-Geisser Test
IOP=Intra-Ocular pressure; BCVA= Best Corrected Visual Acuity

To further assess the safety of RMS, we compared IOP values over time between treated and untreated groups. The IOP measurements at baseline (enrolment) and subsequent time points (1 week, 1 month, 2 months, and 3 months) are presented in table 2. Overall, while both groups exhibited decreases in mean IOP over the 3-month period, both treated groups did not show consistent reduction. Nevertheless, both groups exhibited stability in the IOP and showed no statistically significant changes over time as shown in [Table 2].

Moreover, to further investigate the safety of RMS treatment, we tracked the change in Schirmer change over time in two distinct treatment groups: the Not Treated group and the Treated group over a three-month period. The standard deviation and mean values are summarized in [table 2]. In the non-treated group despite observable fluctuations, the change over time within this group did not achieve statistical significance (p = 0.51). Similarly, in the Treated group the change over time within the Treated group did not show statistical significance (p = 0.417).

When comparing the change over time between the two groups, no statistically significant difference emerged (p > 0.05). Both the Not Treated and Treated groups displayed variable Schirmer test values over the study period, but with no statistically significant change between the groups.

For the evaluation of efficacy, over the span of 3 months we measured the National Eye Institute (NEI) for each eye at different time points. We have evaluated the total scores across different time points for both the treated and non-treated groups. Table 3 provides a summary of the mean NEI total scores along with corresponding sample sizes and the results of statistical comparisons as shown in [Table 2].

**Table 3.**
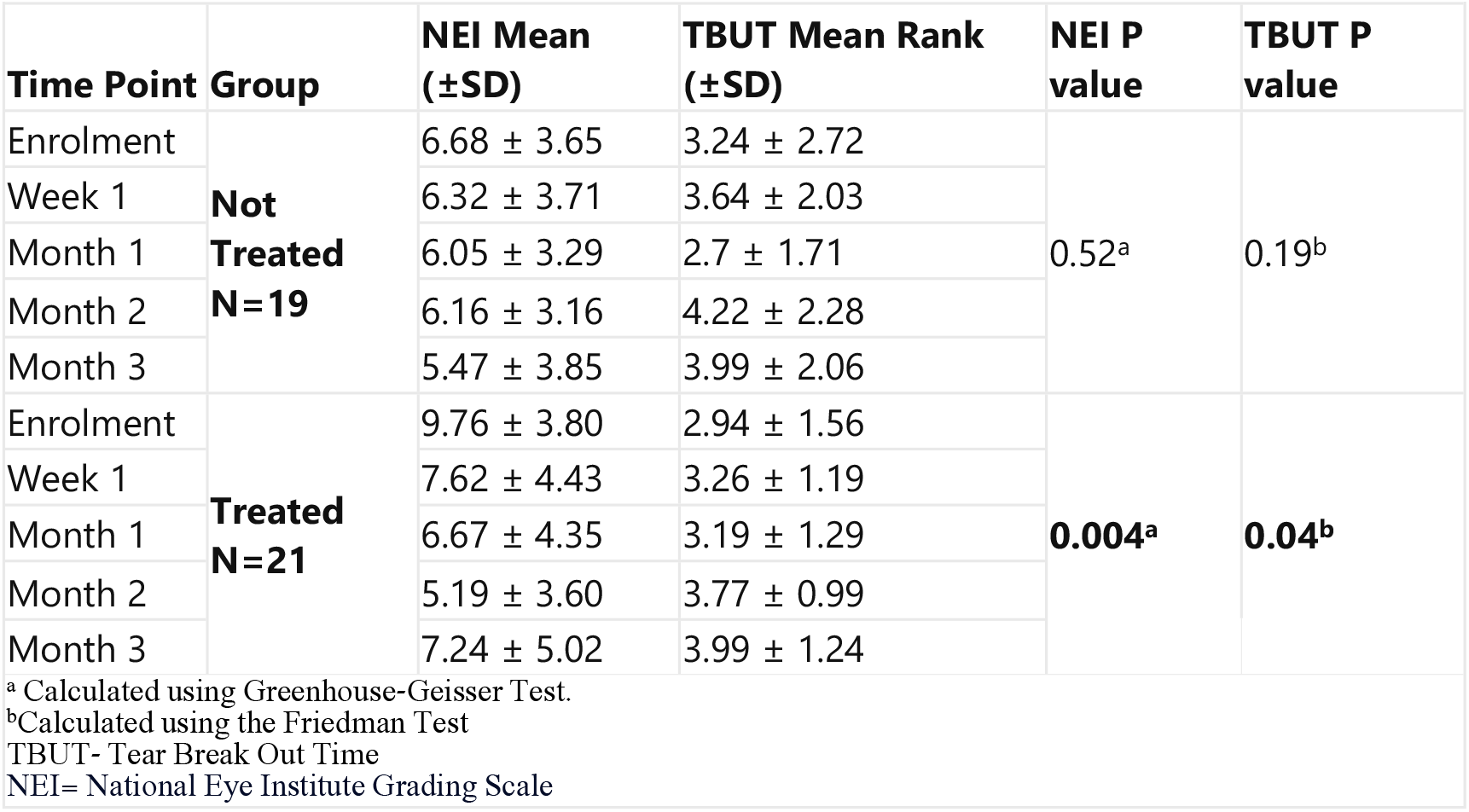
Comparative Analysis of NEI and TBUT Scores in treatment Groups Over Time.

|  |  |  |  |  |  |
| --- | --- | --- | --- | --- | --- |
| Time Point | Group | NEI Mean (±SD) | TBUT Mean Rank (±SD) | NEI P value | TBUT P value |
| Enrolment | Not Treated<br>N=19 | 6.68 ± 3.65 | 3.24 ± 2.72 | 0.52<sup>a</sup> | 0.19<sup>b</sup> |
| Week 1 |  | 6.32 ± 3.71 | 3.64 ± 2.03 |  |  |
| Month 1 |  | 6.05 ± 3.29 | 2.7 ± 1.71 |  |  |
| Month 2 |  | 6.16 ± 3.16 | 4.22 ± 2.28 |  |  |
| Month 3 |  | 5.47 ± 3.85 | 3.99 ± 2.06 |  |  |
| Enrolment | Treated<br>N=21 | 9.76 ± 3.80 | 2.94 ± 1.56 | 0.004<sup>a</sup> | 0.04<sup>b</sup> |
| Week 1 |  | 7.62 ± 4.43 | 3.26 ± 1.19 |  |  |
| Month 1 |  | 6.67 ± 4.35 | 3.19 ± 1.29 |  |  |
| Month 2 |  | 5.19 ± 3.60 | 3.77 ± 0.99 |  |  |
| Month 3 |  | 7.24 ± 5.02 | 3.99 ± 1.24 |  |  |
| <sup>a</sup> Calculated using Greenhouse-Geisser Test. |  |  |  |  |  |
| <sup>b</sup>Calculated using the Friedman Test |  |  |  |  |  |
| TBUT- Tear Break Out Time |  |  |  |  |  |
| NEI= National Eye Institute Grading Scale |  |  |  |  |  |

In the non-treated eyes group, the mean NEI scores displayed no statistically significant change across different time intervals. In contrast, the treated group exhibited a significant pattern of change). The statistical significance of these changes was evaluated and showed revealing a significant difference in the treated group’s NEI total scores over time (P = 0.004). However, the NEI total scores in the non-treated group did not show significant changes over time (P > 0.05). These scores are presented in [Fig 1].

**Fig 1:**
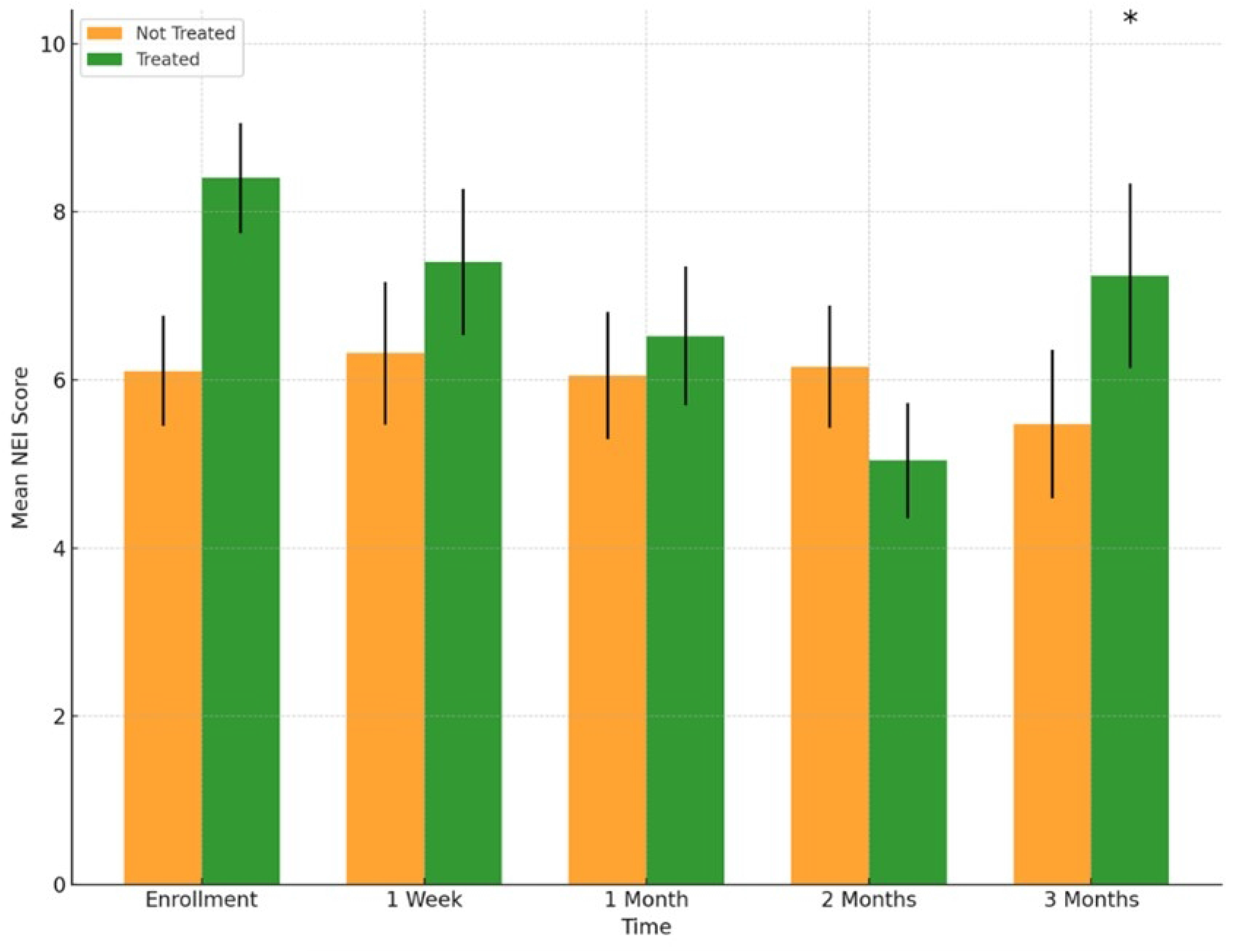
Comparative Assessment of NEI Scores Over Time. This graph illustrates the changes in the National Eye Institute (NEI) Total Scores over a period of three months, comparing groups that were treated (Green) and not treated (Orange). Each bar represents the mean ±SE NEI Total Score at five time points: Enrollment, 1 Week, 1 Month, 2 Months, and 3 Months. The black asterisks represent the statistical significance in the differences observed.

Efficacy of the RMS was also assessed by Tear Break-Up Time (TBUT) changes over a three-month period in the two groups: a Non-Treated Group and a Treated Group. [Table 3].

For the non-treated group, no statistically significant changes in TBUT scores across the designated time intervals (P=0.198). Contrarily, TBUT scores for the treated group showed an improvement in the period tested with statistical significance (p=0.045). These changes with the pattern of change of the testing period are showed in [Fig 2].

**Fig 2:**
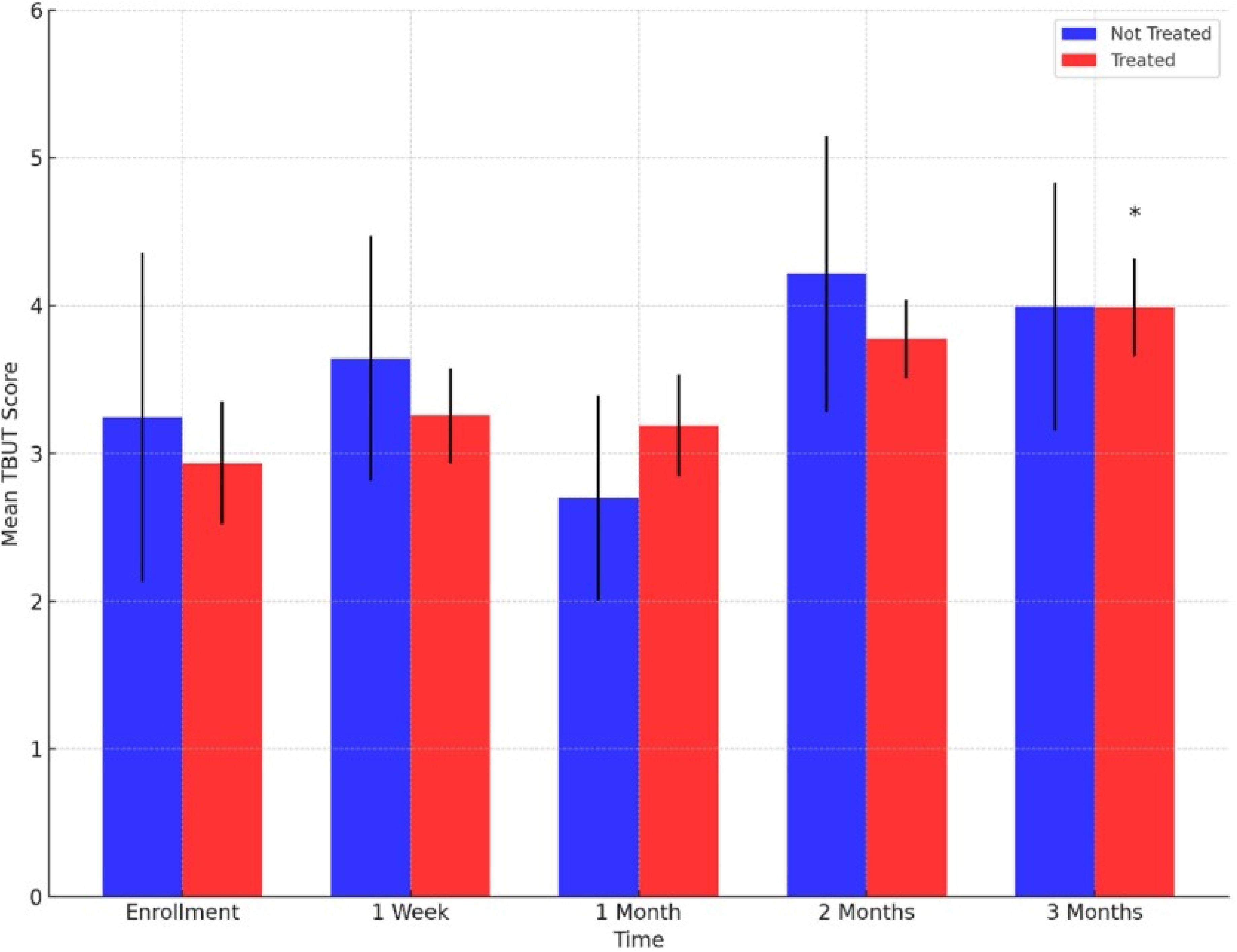
Comparative Assessment of Tear Break Out Time Between Treatment Groups. This figure illustrates the temporal progression of Tear Break-Up Time (TBUT) scores across five different time points. The graph is comparing groups that were treated (red) and not treated (Blue). Each bar represents the mean ±SE TBUT score. The black asterisks represent the statistical significance in the differences observed.

In addition, an evaluation of the Subjective assessment scores was performed for both the non-treated and the treated groups. Over a corresponding 3-month period and matching time points, the Subjective score for each patient was measured [Table 4].

**Table 4.** Subjective assessment Score Change Over Time by Treatment Group.

| Treatment Group | Time | Score Mean (±SD) | P<sup>a</sup> |
| --- | --- | --- | --- |
| **Not Treated** | Enrolment Score | 12.50 (±1.55) | **0.008** |
| | 1 week score | 8.27 (±1.36) | |
| | 1 month score | 8.09 (±1.62) | |
| | 2-month score | 7.27 (±1.61) | |
| | 3-month score | 8.45 (±1.45) | |
| **Treated** | Enrolment Score | 7.42 (±1.26) | **0.007** |
| | 1 week score | 5.95 (±1.07) | |
| | 1 month score | 5.65 (±1.23) | |
| | 2-month score | 6.65 (±1.38) | |
| | 3-month score | 5.81 (±1.45) | |

<sup>a</sup>Calculated using the Friedman Test

The Subjective assessment scores exhibited notable changes throughout the study. Both the treated and non-treated groups had statistically significant changes (P<0.001 and P<0.005 respectively). These changes with the pattern of change of the testing period are showed in [Fig 3]

**Fig 3.**
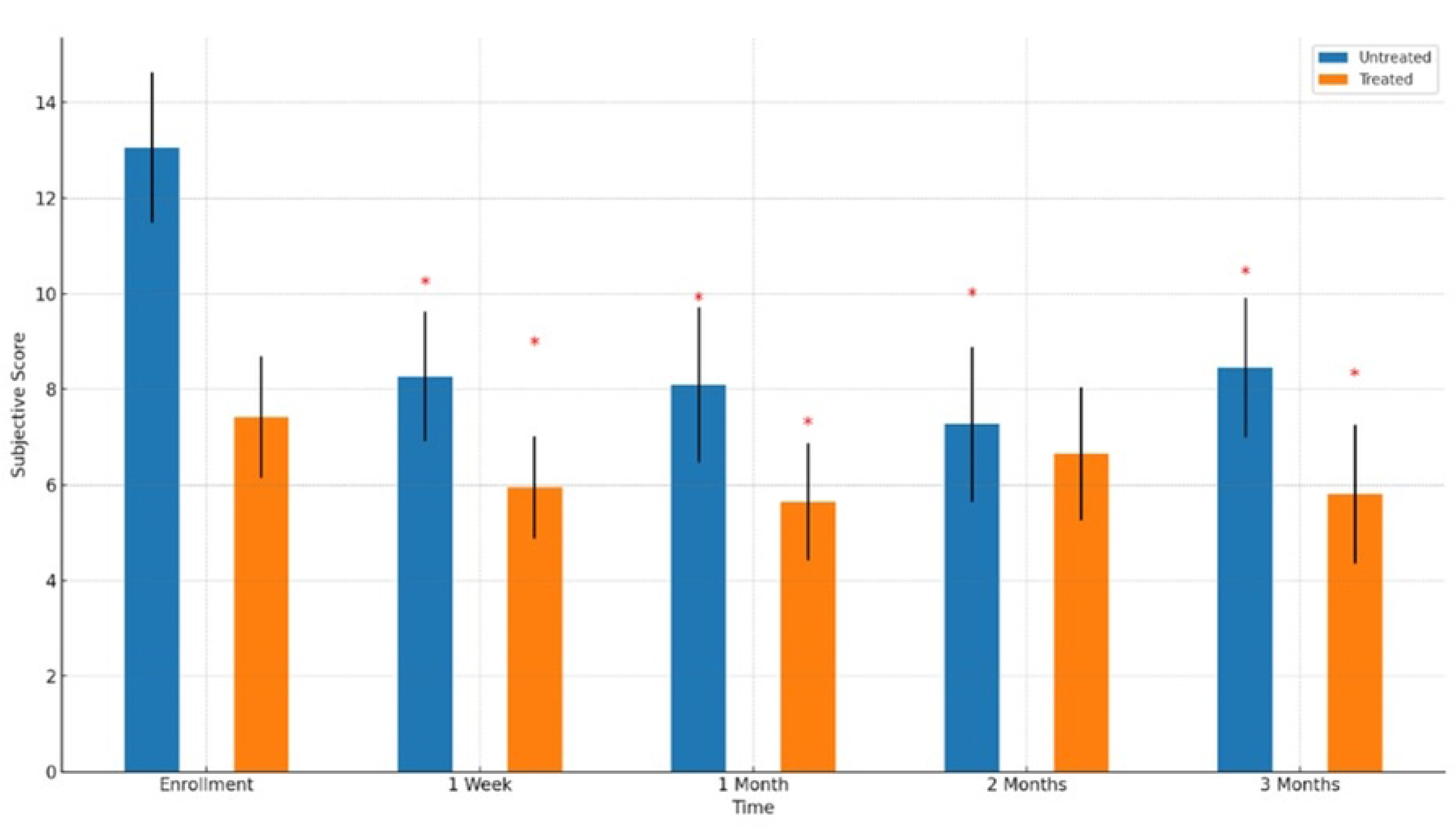
Subjective Assessment Score Change Over Time by Treatment Group. This graph illustrates the changes in the National Eye Institute (NEI) Total Scores over a period of three months, comparing groups that were treated (Orange) and not treated (Blue). Each bar represents the mean ±SE NEI Total Score at five time points: Enrollment, 1 Week, 1 Month, 2 Months, and 3 Months. The red asterisks represent the statistical significance in the differences observed.

## Discussion

In the present study, the safety and initial efficacy of repetitive magnetic stimulation (RMS) as a pioneering, first-in-human therapeutic intervention for dry eye syndrome was explored. This study encompassed 22 adult subjects diagnosed with moderate to severe dry eye syndrome, who underwent treatment utilizing the VIVEYE - Ocular Magnetic Neurostimulation System Ver 1.0. The primary endpoints of this research were the safety, tolerability, and preliminary effectiveness of the intervention. Our findings indicate that RMS is a safe and well-tolerated modality, which resulted in significant amelioration of dry eye symptoms, as evidenced by improved fluorescein staining scores and reduced patient-reported ocular discomfort, thereby underscoring the innovative potential of this treatment in managing dry eye disease.

Neurostimulation, as indicated in prior animal research, influences epithelial cells via the activation of trigeminal nerve endings [21][22][23]. The suggested mechanism for this activation is through secretion of neurotransmitters and neurotrophic factors [23], although the exact nature is unclear. RMS may have therapeutic effect on the parasympathetic innervation of the lacrimal gland and conjunctival goblet cells[24]. In trials involving repetitive transcranial magnetic stimulation, long-term impacts have been linked to alterations in gene and protein expression [25][26][27][28]. Substance P, a trigeminal nerve neuropeptide has been observed to promote cell attachment through E-cadherin and stimulate DNA synthesis and growth in vitro [29][30], leading to epithelial cell proliferation [31] Collectively, these findings suggest that RMS treatment in animal models may offer corneal protection, positioning it as a potential safe therapeutic option for patients with exposure keratopathy [23]

Our study, drawing on precedents from prior research (28), indicates that the therapeutic efficacy of Repetitive Magnetic Stimulation (RMS) in alleviating Dry Eye Syndrome primarily operates through the proliferation of epithelial cells. Consistent Schirmer test scores observed across the study period suggest that the observed improvements in both subjective and objective measures are not attributable to alterations in tear secretion. Notably, significant modifications were detected in parameters indicative of epithelial transformation, including National Eye Institute (NEI) scores and Tear Film Break-Up Time (TBUT), lending further support to the proposition that RMS promotes epithelial cell changes and proliferation. This aligns with the hypothesized mechanism of RMS action involving direct stimulation of trigeminal nerve fibers, which leads to the release of neuromodulators from sensory nerve endings, thereby directly stimulating corneal epithelial growth.

Several previous studies have investigated the use of different forms of neurostimulation in humans for the treatment of dry eye disease. A study that investigated the use of transcutaneous electrical nerve stimulation (TENS) [32] found that this treatment improved symptoms of dry eye, including ocular discomfort and tear film instability. Similarly, the use intranasal tear stimulator (ITS) [33] was reported to improved symptoms of dry eye. Moreover, in animals, treatment with electrical stimulation of the lacrimal gland (LNS) and afferent nerves for enhanced tear secretion [34] also found to improve symptoms of dry eye.

All these studies, that were developed to treat dry eye signs and symptoms and are based on electric stimulation, including the current one, suggest that neurostimulation may be a promising treatment option for dry eye disease. However, it is worth noting that these studies used different forms of neurostimulation (TENS, LNS, ITS, RMS) and different stimulation parameters. It is therefore difficult to directly compare the results of these treatments and further research is needed to determine the optimal form and parameters of neurostimulation for the treatment of dry eye disease.

Contrary to the methodologies referenced in preceding studies, our investigation showcases distinct advantages that potentially augment treatment outcomes and patient compliance. For instance, while previous studies on Transcutaneous Electrical Nerve Stimulation (TENS) required a regimen of 20 sessions spread over five sessions per week (29), our protocol involved a mere four treatment sessions. Furthermore, unlike studies focusing on intranasal tear stimulation (30), our method is entirely non-invasive, thereby circumventing the adverse effects associated with invasive procedures, such as epistaxis. These distinctions underscore the benefits of utilizing repetitive magnetic stimulation over the methods employed in these prior studies, offering a non-invasive and more manageable treatment approach. Consequently, these characteristics are likely to enhance patient adherence and overall satisfaction with the treatment regimen, underscoring the potential for improved clinical outcomes and patient experiences.

The current study is subject to several limitations that might impact the extrapolation of its findings. Firstly, the relatively small sample size, a consequence of the high costs associated with conducting the trial, may restrict the generalizability of the results. Additionally, the cohort consisted exclusively of individuals presenting with moderate to severe dry eye syndrome, suggesting that the observed efficacy of repetitive magnetic stimulation (RMS) may not extend to those with milder forms of the condition. The potential for bias is further compounded by the study’s open-label design, where some participants were allocated to a placebo group, and others received active treatment in both eyes. Variability in the subjective assessment questionnaires, despite efforts to maintain consistency in scoring and analysis, introduces another layer of complexity that could potentially lead to statistical inaccuracies and fail to accurately reflect the progression of symptoms over time.

Despite these methodological constraints, our study identified significant improvements in signs and symptoms associated with dry eye disease among the participants. These key findings highlight the potential of RMS as an effective intervention for reducing symptoms of dry eye syndrome, underscoring its promise despite the noted limitations.

This study presents an innovative approach to the management of Dry Eye Disease (DED), introducing a non-invasive, cost-effective, and efficient therapeutic method. Our findings indicate that Repetitive Magnetic Stimulation (RMS) promotes epithelial healing and elicits notable improvements in both objective dry eye parameters and subjective patient experiences, with favorable tolerance levels reported. To our knowledge, the application of RMS in DED treatment represents a novel exploration not previously documented in the literature. This contrasts with its use in psychiatric conditions, such as obsessive-compulsive disorder (OCD), where treatment typically necessitates multiple sessions per week. Our protocol, however, implemented RMS in a singular session format. This initial evidence supports the potential for further investigation and broader application of this technology, including modifications to stimulus frequency, timing, and an increased number of treatment sessions, which may unveil greater benefits in DED management.

Furthermore, corroborating evidence from animal studies involving rabbits suggests RMS as an efficacious intervention for corneal protection and a safe, efficient treatment for patients with exposure keratopathy. Its non-invasive, pain-free nature, coupled with rapid and significant therapeutic outcomes, positions RMS as a viable alternative for patients unresponsive to conventional therapies. The infrequent need for treatment sessions, spaced months apart, enhances its cost-effectiveness and convenience, particularly by obviating the daily use of eye drops. The simplicity and minimal maintenance requirements of this therapy make it especially appealing for patients reluctant to undergo invasive procedures or those with contraindications to existing treatments. Moreover, the technology’s ease of use extends its applicability to a broad range of healthcare providers, including office staff, ophthalmologists, optometrists, and nursing staff.

While our findings are encouraging, further research is imperative to corroborate these results comprehensively and assess the long-term impacts of magnetic stimulation on dry eye disease treatment.

### Conclusions

This study introduces a groundbreaking method by applying repetitive magnetic stimulation (RMS) as a non-invasive treatment for dry eye syndrome. The outcomes underscore RMS’s safety and its potential efficacy in promoting tear film stability, mitigating corneal damage, and alleviating patient-reported symptoms, all without negative effects on intraocular pressure, visual acuity, or tear secretion. These findings suggest the feasibility of incorporating RMS into the treatment repertoire for dry eye disease, indicating a potential paradigm shift towards non-pharmacological therapeutic approaches. Nonetheless, additional research is required to delineate the long-term efficacy of this treatment and to refine the treatment protocols, heralding a novel avenue in ophthalmic treatment strategies.

## Acknowledgements

This paper was part of the requirements for Shimon Perelman’s MD title from the Hebrew University, Jerusalem, Israel. This research received no specific grant from any agency.

## Data Availability

The data set generated during the current study is available from the corresponding authors on reasonable request.

## References

[1] J. Gayton, “Etiology, prevalence, and treatment of dry eye disease,” Clin Ophthalmol., vol. 3, pp. 405–412, 2009.

[2] D. F. Courtin R, Pereira B, Naughton G, Chamoux A, Chiambaretta F, Lanhers C, “Prevalence of dry eye disease in visual display terminal workers: a systematic review and meta-analysis,” BMJ Open, vol. 6, no. 1, 2016, doi: 10.1136/bmjopen-2015-009675.

[3] A. Galor and M. Miami, “Painful Dry Eye Symptoms : A Nerve Problem or a Tear Problem ?,” Ophthalmology, vol. 126, no. 5, pp. 648–651, 2019, doi: 10.1016/j.ophtha.2019.01.028.

[4] I. Sher et al., “Repetitive magnetic stimulation protects corneal epithelium in a rabbit model of short-term exposure keratopathy,” Ocular Surface, 2019, doi: 10.1016/j.jtos.2019.09.009.

[5] G. Dieckmann, F. Fregni, and P. Hamrah, “Neurostimulation in dry eye disease—past, present, and future,” Ocular Surface, vol. 17, no. 1, pp. 20–2, 2019, doi: 10.1016/j.jtos.2018.11.002.

[6] J. D. Sheppard et al., Characterization of tear production in subjects with dry eye disease during intranasal tear neurostimulation: Results from two pivotal clinical trials. Elsevier Inc. doi: 10.1016/j.jtos.2018.11.009.

[7] L. Carmi, U. Alyagon, N. Barnea-ygael, J. Zohar, R. Dar, and A. Zangen, “Clinical and electrophysiological outcomes of deep TMS over the medial prefrontal and anterior cingulate cortices in OCD patients,” Brain Stimul, vol. 11, no. 1, pp. 158–165, 2018, doi: 10.1016/j.brs.2017.09.004.

[8] S. Fiocchi, M. Longhi, P. Ravazzani, Y. Roth, A. Zangen, and M. Parazzini, “Modelling of the Electric Field Distribution in Deep Transcranial Magnetic Stimulation in the Adolescence, in the Adulthood, and in the Old Age,” Comput Math Methods Med., 2016.

[9] B. J. Seewoo, K. W. Feindel, S. J. Etherington, and J. Rodger, “Frequency-specific effects of low-intensity rTMS can persist for up to 2 weeks post-stimulation: A longitudinal rs-fMRI / MRS study in rats,” Brain Stimul, no. xxxx, 2019, doi: 10.1016/j.brs.2019.06.028.

[10] “NCT03012698 @ clinicaltrials.gov.”

[11] A. Behrens et al., “Dysfunctional tear syndrome: a Delphi approach to treatment recommendations.,” Cornea, vol. 25, no. 8, pp. 900–907, 2006.

[12] P. Versura, V. Profazio, and E. C. Campos, “Performance of tear osmolarity compared to previous diagnostic tests for dry eye diseases.,” Curr Eye Res, vol. 35, no. 7, pp. 553–564, 2010, doi: 10.3109/02713683.2010.484557.

[13] I. Sher et al., “Multimodal Assessment of Corneal Erosions Using Optical Coherence Tomography and Automated Grading of Fluorescein Staining in a Rabbit Dry Eye Model,” Transl Vis Sci Technol, vol. 28;8, no. 1, p. :27, 2019.

[14] D. F. Sweeney, T. J. Millar, and S. R. Raju, “Tear film stability : A review,” Exp Eye Res, vol. 117, pp. 28–38, 2013, doi: 10.1016/j.exer.2013.08.010.

[15] “The epidemiology of dry eye disease: report of the Epidemiology Subcommittee of the International Dry Eye WorkShop (2007),” Ocul Surf., vol. 5, no. 2, pp. 93–107, 2007, doi: 10.1016/S1542-0124(12)70082-4.

[16] S. J. Bron AJ, Evans VE, “Grading Of Corneal and Conjunctival Staining in the Context of Other Dry Eye Tests,” Cornea, vol. 22, no. 7, pp. 640–650, 2003.

[17] P. IK. Chun YS, Yoon WB, Kim KG, “Objective assessment of corneal staining using digital image analysis.,” Invest Ophthalmol Vis Sci, vol. 55, no. 12, pp. 7896–903, 2014, doi: 10.1167/iovs.14-15618.

[18] W. Ngo, P. Situ, N. Keir, D. Korb, C. Blackie, and T. Simpson, “Psychometric Properties and Validation of the Standard Patient Evaluation of Eye Dryness Questionnaire,” Cornea, vol. 32, no. 9, pp. 1204–1210, Sep. 2013, doi: 10.1097/ICO.0b013e318294b0c0.

[19] R. D. Hays et al., “Assessment of the Psychometric Properties of a Questionnaire Assessing Patient-Reported Outcomes With Laser In Situ Keratomileusis (PROWL),” JAMA Ophthalmol, vol. 135, no. 1, p. 3, Jan. 2017, doi: 10.1001/jamaophthalmol.2016.4597.

[20] A. Bond and M. Lader, “The use of analogue scales in rating subjective feelings,” British Journal of Medical Psychology, vol. 47, no. 3, pp. 211–218, Sep. 1974, doi: 10.1111/j.2044-8341.1974.tb02285.x.

[21] R. W. Beuerman and B. Schimmelpfennig, “Sensory denervation of the rabbit cornea affects epithelial properties,” Exp Neurol, vol. 69, no. 1, pp. 196–201, Jul. 1980, doi: 10.1016/0014-4886(80)90154-5.

[22] D. J. Oswald et al., “Communication between Corneal Epithelial Cells and Trigeminal Neurons Is Facilitated by Purinergic (P2) and Glutamatergic Receptors,” PLoS One, vol. 7, no. 9, p. e44574, Sep. 2012, doi: 10.1371/journal.pone.0044574.

[23] I. Sher et al., “Repetitive magnetic stimulation protects corneal epithelium in a rabbit model of short-term exposure keratopathy,” Ocular Surface, vol. 18, no. 1, pp. 64–73, Jan. 2020, doi: 10.1016/j.jtos.2019.09.009.

[24] H. Toshida, D. H. Nguyen, R. W. Beuerman, and A. Murakami, “Evaluation of Novel Dry Eye Model: Preganglionic Parasympathetic Denervation in Rabbit,” Investigative Opthalmology & Visual Science, vol. 48, no. 10, p. 4468, Oct. 2007, doi: 10.1167/iovs.06-1486.

[25] S. fang Feng, T. yao Shi, Fan-Yang, W. ning Wang, Y. chun Chen, and Q. rong Tan, “Long-lasting effects of chronic rTMS to treat chronic rodent model of depression,” Behavioural Brain Research, vol. 232, no. 1, pp. 245–251, Jun. 2012, doi: 10.1016/J.BBR.2012.04.019.

[26] H.-Y. Wang et al., “Repetitive Transcranial Magnetic Stimulation Enhances BDNF-TrkB Signaling in Both Brain and Lymphocyte,” Journal of Neuroscience, vol. 31, no. 30, pp. 11044–11054, Jul. 2011, doi: 10.1523/JNEUROSCI.2125-11.2011.

[27] K. Kudo et al., “Repetitive transcranial magnetic stimulation induces kf-1 expression in the rat brain,” Life Sci, vol. 76, no. 21, pp. 2421–2429, Apr. 2005, doi: 10.1016/J.LFS.2004.10.046.

[28] J. Hellmann et al., “Repetitive magnetic stimulation of human-derived neuron-like cells activates cAMP-CREB pathway,” Eur Arch Psychiatry Clin Neurosci, vol. 262, pp. 87–91, 2012.

[29] T. W. Reid, C. J. Murphy, C. K. Iwahashi, B. A. Foster, and M. J. Mannis, “Stimulation of epithelial cell growth by the neuropeptide substance P,” J Cell Biochem, vol. 52, no. 4, pp. 476–485, Aug. 1993, doi: 10.1002/jcb.240520411.

[30] K. Araki-Sasaki et al., “Substance P-induced cadherin expression and its signal transduction in a cloned human corneal epithelial cell line,” J Cell Physiol, vol. 182, no. 2, pp. 189–195, Feb. 2000, doi: 10.1002/(SICI)1097-4652(200002)182:2<189::AID-JCP7>3.0.CO;2-9.

[31] J. Garcia-Hirschfeld, L. G. Lopez-Briones, and C. Belmonte, “Neurotrophic Influences on Corneal Epithelial Cells,” Exp Eye Res, vol. 59, no. 5, pp. 597–605, Nov. 1994, doi: 10.1006/EXER.1994.1145.

[32] M.-M. Cai and J. Zhang, “Effectiveness of transcutaneous electrical stimulation combined with artificial tears for the treatment of dry eye: A randomized controlled trial,” Exp Ther Med, vol. 20, no. 6, 2020, doi: 10.3892/etm.2020.9305.

[33] J. K. Park, S. Cremers, and A. L. Kossler, “Neurostimulation for tear production,” Curr Opin Ophthalmol, vol. 30, no. 5, pp. 386–394, Sep. 2019, doi: 10.1097/ICU.0000000000000590.

[34] M. Brinton et al., “Electronic enhancement of tear secretion,” J Neural Eng, vol. 13, no. 1, p. 016006, Feb. 2016, doi: 10.1088/1741-2560/13/1/016006.

